# Spectral power and functional connectivity of resting EEG in people exposed to armed conflict and controls

**DOI:** 10.64898/2026.10.15.26365676

**Authors:** Laura Nitola Pulido, Santiago Bonilla, Eduar Herrera, Pablo Reyes, Hernando Santamaría García

## Abstract

Exposure to armed conflict can leave persistent effects on mental health, yet its neurophysiological correlates remain poorly characterized, particularly in populations exposed to prolonged violence outside narrowly defined post traumatic stress disorder samples. Here we examined resting state electroencephalographic activity in 122 Colombian adults, including 74 participants exposed to armed conflict and 48 controls. Eyes closed EEG was analysed for 240 seconds per participant. We quantified absolute and relative spectral power across delta, theta, alpha and beta bands and regional functional connectivity using weighted phase lag index in theta, alpha and beta. Group comparisons were adjusted for age, education, sex and assessment year, with correction across 78 EEG contrasts. Median occipital and global alpha power was lower in exposed participants, although adjusted estimates were close to the null and confidence intervals were broad. Temporal theta relative power was lower in exposed participants at nominal significance; the contrast did not retain statistical significance after Holm correction. No spectral power or regional connectivity contrast met the adjusted significance threshold. Exploratory associations between EEG measures and MoCA performance were modest among participants without scoring flags. These findings characterize resting EEG in a population shaped by prolonged violence and displacement. The confidence intervals leave the magnitude and direction of several group differences uncertain, while the observed alpha and theta patterns identify questions for larger studies integrating more detailed exposure and cognitive measures.

## 1 Introduction

Armed conflict exposes individuals and communities to repeated violence, threat, bereavement, displacement and disruption of social and material conditions across the life course. Its consequences for mental health are well established. A large systematic review estimated that approximately one in five people living in conflict affected settings experience depression, anxiety, post traumatic stress disorder or other mental disorders at any point in time [1]. Severe and repeated trauma has also been associated with persistent alterations in systems supporting stress regulation, emotion, memory and threat processing [2]. Yet the effects of armed conflict on brain health remain much less understood than its psychiatric consequences. Studies using neuroimaging, electrophysiology and peripheral biomarkers remain comparatively sparse, particularly in populations living with the cumulative social consequences of conflict. Understanding these biological correlates may help characterize how lived experiences of violence become embedded in neural function across adulthood.

Available evidence suggests that severe trauma and post traumatic stress disorder involve distributed changes across neural systems. Neuroimaging studies have repeatedly implicated the amygdala, hippocampus, anterior cingulate and prefrontal regions, together with altered interactions among networks supporting salience, memory and emotional regulation [2]. Electrophysiological studies provide a complementary window into these processes. A systematic review identified recurrent associations between post traumatic stress symptoms and alpha rhythms and event related potentials, although findings varied substantially across studies [3]. Resting EEG studies have reported altered theta and alpha power and connectivity in PTSD, including increased frontal and parietal theta activity, altered alpha connectivity and reduced efficiency of large scale cortical networks across theta and beta frequencies [4, 5]. These studies suggest that trauma related states can affect the spectral and temporal organization of neural activity. Most evidence, however, comes from clinically selected PTSD samples, military populations or specific trauma groups, leaving the neurophysiological consequences of broader armed conflict exposure less clearly defined.

Colombia provides a particularly important setting in which to address this gap. The country has experienced more than seven decades of recurrent political violence and more than six decades of armed conflict, accompanied by mass forced displacement, poverty, social exclusion and persistent inequalities in access to care. Recent work has documented substantial burdens of PTSD, depression, anxiety and somatic symptoms among people affected by the conflict, including internally displaced populations [6]. The consequences extend beyond psychiatric symptoms, and recent perspectives have argued that violence, displacement and social adversity may be contributing to an emerging brain health crisis in the country [7]. Despite this context, neurobiological studies remain limited. Very little is known about resting EEG signatures of conflict exposure in Colombian populations, and evidence is particularly scarce for frequency specific spectral alterations and functional connectivity. This gap also limits understanding of whether neural patterns identified in PTSD studies generalize to people exposed to prolonged conflict with heterogeneous clinical presentations.

Here we examined eyes closed resting EEG in adults exposed to armed conflict and controls. We standardized the analysed duration to 240 seconds per participant and quantified absolute and relative power across delta, theta, alpha and beta bands in frontal, central, parietal, occipital and temporal sensor cores. Functional connectivity was estimated using weighted phase lag index across theta, alpha and beta frequencies. Group differences were adjusted for age, education, sex and assessment year, and statistical inference accounted for multiple comparisons across the complete EEG analysis. We also examined the robustness of alpha findings across reference schemes and assessment year and explored associations between selected EEG measures and global cognition measured with the MoCA.

We hypothesized that exposure to armed conflict would be associated with altered resting spectral organization and functional connectivity, with the strongest differences expected in theta and alpha frequencies given previous trauma related EEG findings. We further expected that EEG alterations would extend across regional sensor networks and show associations with cognitive performance. More broadly, we sought to determine whether prolonged exposure to armed conflict is accompanied by measurable resting state neurophysiological signatures in a Colombian population shaped by violence and its social consequences.

## 2 Methods

### 2.1 Study objective

To compare spectral power and functional connectivity between people exposed to armed conflict and controls using the same total EEG duration and common regional sensor cores. MoCA is integrated through a scoring review and descriptive exploration of its relationships with EEG to support a more complete cognitive comparison.

### 2.2 EEG selection and preprocessing

Processing used nominal BioSemi 64-channel EEG identities. F4 and P4 were excluded consistently, as were inputs with unknown locations, including EXG1 and EXG2. Initial channel screening used consecutive ten-second blocks of the original recording. Channels were excluded for non-finite samples, calibration-limit saturation, a median block standard deviation below 2 *µ*V, or persistently extreme amplitude. The last criterion required a median peak-to-peak amplitude above 500 *µ*V, at least five times the recording’s median across EEG channels, together with peak-to-peak amplitude above 500 *µ*V in at least 20% of blocks. Saturation was flagged when at least 0.1% of a block’s samples were within one digital calibration step of either physical limit.

The first and last ten seconds were removed, and the continuous signal was resampled to 512 Hz using polyphase FIR resampling with anti-aliasing. Signals were referenced to the average of usable EEG channels. Filtering used Butterworth IIR designs implemented as second-order sections: a 60 Hz notch with design order parameter 2, followed by a 0.5-45 Hz band-pass with design order parameter 4. Both filters were applied forward and backward for zero-phase filtering [8]. A further five seconds were discarded at each end after filtering to reduce boundary effects. Preprocessing used MNE-Python 1.13.2.

The remaining signal was divided into consecutive, non-overlapping four-second epochs without baseline correction. A channel-epoch was flagged for non-finite values, peak-to-peak amplitude above 250 *µ*V, standard deviation above 80 or below 0.05 *µ*V, or a maximum absolute first difference multiplied by the sampling rate above 25,600 *µ*V/s. Channels flagged in at least half of the candidate epochs were removed and the average reference was recalculated. An epoch was accepted only when all remaining channels met these criteria. At least 48 usable channels and 60 accepted epochs were required. ICA and interpolation were not applied.

For each participant, 60 epochs were selected at evenly spaced positions in the chronologically ordered set of accepted epochs, standardising the analysed duration to 240 seconds. At 512 Hz, each four-second epoch contains 2,048 samples and yields a Fourier frequency spacing of 0.25 Hz. This duration provided a common frequency grid for band integration while permitting the exclusion of brief segments that failed the signal criteria. Using the same number and length of epochs also kept the amount of data contributing to power and connectivity estimates consistent across participants.

### 2.3 Regional sensor cores

Figure 1 locates the 14 positions used in the regional sensor cores and connectivity matrices. The frontal, central, parietal and occipital cores each contain three positions; the temporal core contains two. The global core is the union of all 14 positions.

**Figure 1.**
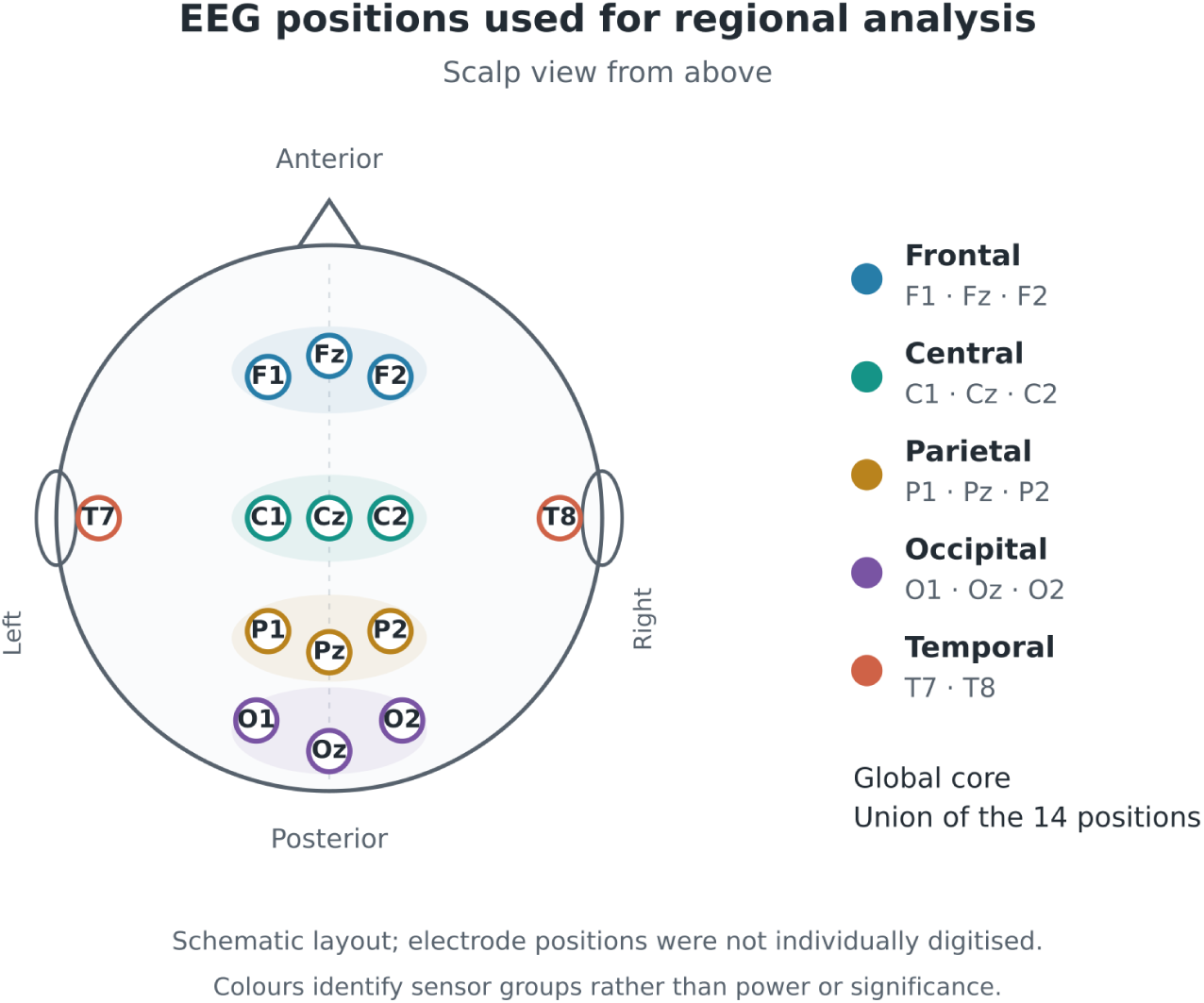
Regional EEG sensor cores viewed from above. Colours identify sensor groups and do not encode power, connectivity or statistical significance.

The layout is schematic and does not use individually digitised electrode positions. These cores describe scalp measurements rather than complete lobes or localised cortical sources. Table 2 and Figure 3 show their availability by study group.

**Table 1.**
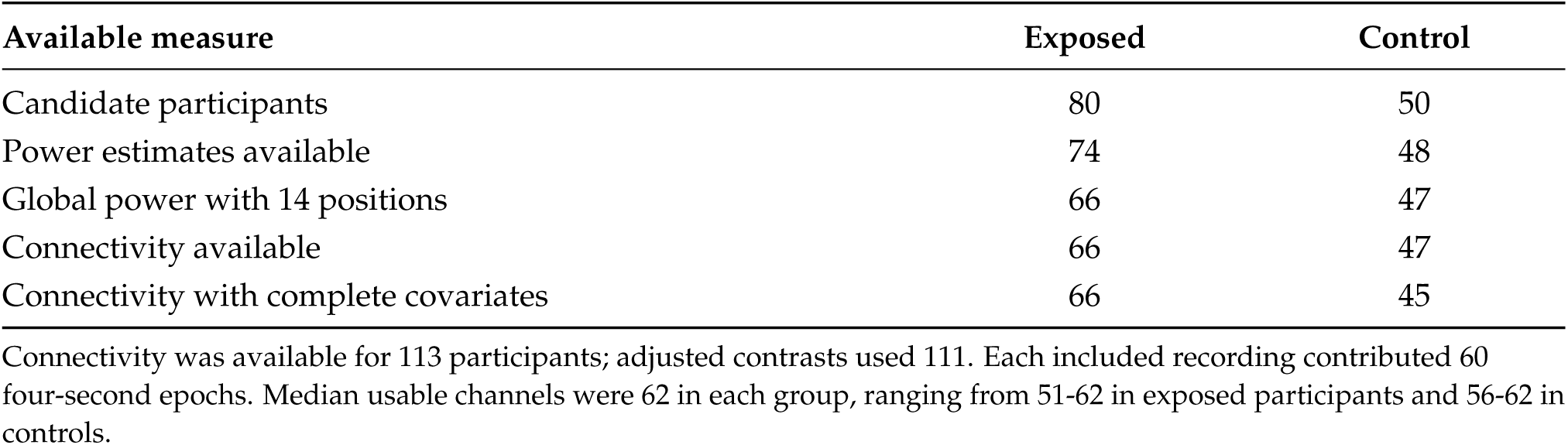
Availability of recordings and measures by study group.

**Table 2.** Regional sensor cores and participants with all required positions.

| Sensor core | Required channels | Exposed | Control |
| --- | --- | --- | --- |
| Frontal | F1 Fz F2 | 72 | 47 |
| Central | C1 Cz C2 | 73 | 48 |
| Parietal | P1 Pz P2 | 71 | 48 |
| Occipital | O1 Oz O2 | 70 | 48 |
| Temporal | T7 T8 | 73 | 48 |
| Global 14 | Union of the 14 channels | 66 | 47 |

**Table 3.** Characteristics of participants with EEG power estimates.

| Characteristic | Exposed | Control |
| --- | --- | --- |
| Age in years | 40.99 (13.06); n=74 | 34.41 (12.01); n=46 |
| Education in years | 9.00 (4.01); n=74 | 12.15 (2.36); n=46 |
| Sex code 1 | 22 (29.7 %) | 19 (39.6 %) |
| Sex code 2 | 52 (70.3 %) | 27 (56.2 %) |
| Sex code missing | 0 | 2 |
| Assessment year missing | 0 | 2 |

Age and education: mean (SD) and available sample size. Categorical values: count and percentage within the EEG group. Sex codes have unconfirmed meanings. Two controls have incomplete covariates.

### 2.4 Spectral power

Power spectral density (PSD) was estimated as the mean of Hann-windowed periodograms across the 60 epochs, using SciPy 1.18.1. Each four-second epoch was demeaned before applying the window, with a 2,048-point Fourier transform, no zero-padding and no overlap. Frequency bins were spaced 0.25 Hz apart. The bands were delta (1-4 Hz), theta (4-8 Hz), alpha (8-13 Hz) and beta (13-30 Hz). Trapezoidal PSD integration yielded absolute band power in *µ*V^2^.

For each core, PSD was first averaged across its positions and band powers were then calculated. Relative power is the percentage of band power within total 1-30 Hz power for that same core. A core was analysed only when all its positions were available. Relative power describes spectral composition; a change can also reflect changes in other bands.

### 2.5 Functional connectivity

Functional connectivity was quantified using conventional weighted phase-lag index (wPLI), which weights phase-lag contributions by the magnitude of the imaginary cross-spectrum [9]. It was estimated across the same 60 epochs in Fourier mode with Hann windows, using MNE-Connectivity and a reference averaged over Fz/Cz/Pz/Oz. The conventional estimator, rather than the debiased squared estimator, was used. Frequency-specific wPLI values were averaged across 4-7.75, 8-12.75 and 13-30 Hz for theta, alpha and beta, respectively, avoiding duplicated boundary frequencies. There were 91 sensor pairs per participant. Regional connectivity was the mean of the pairs linking two cores, yielding ten regional pairs per band and 30 group contrasts. These maps represent sensor relationships rather than anatomical connections or causal direction.

### 2.6 Adjusted comparisons and sensitivity analyses

The participant was the inferential unit. Linear models included a single exposed/control indicator, with age and education as continuous variables and sex and assessment year as categorical variables. Absolute power was modelled on the base-10 logarithmic scale; relative power was expressed as a percentage and connectivity as wPLI. HC3 robust covariance was used, with confidence intervals (CIs) based on the t distribution. At least ten complete cases per group and ten residual degrees of freedom were required.

Effects are reported as exposed/control ratios of conditional geometric means for absolute power, and exposed-minus-control differences for relative power and wPLI. Holm correction was applied within each family and across the 78 EEG contrasts. The appendices also report Holm values for the planned set of 82 contrasts, including four pending cognitive models entered with p=1; these are not treated as fitted models. The inferential threshold was a Holm-adjusted p<0.05. The 95% CIs are nominal rather than simultaneous. Occipital and global alpha sensitivity was examined using a fixed four-position reference and restriction to the common assessment year, 2025.

### 2.7 MoCA scoring review and descriptive EEG associations

The Montreal Cognitive Assessment (MoCA) is a brief cognitive screening instrument [10]. The full version has a maximum of 30 and allows an education point under specific conditions; the applicable rule must be confirmed for the version used.^1^ Original totals were retained, with domain sums used to guide review. Scores were used as continuous measures; no diagnostic classification was made from a screening cutoff.

Relationships between total MoCA and global 14-position theta/alpha, relative occipital alpha and relative frontal alpha were recalculated. Each relationship included participants without scoring flags and with complete EEG measures and covariates (Table 8).

Residuals of the EEG and MoCA ranks were correlated after adjustment for ranked age and education and categorical sex, assessment year and group. Ties received average ranks. Figure 11 shows the participant residuals in rank units rather than clinical-score units.

### 2.8 Illustration of preprocessed EEG

Figure 2 presents a real four-second epoch selected during processing from a control participant recorded at rest with eyes closed. The five channels illustrate the preprocessed signal in the time domain.

**Figure 2.**
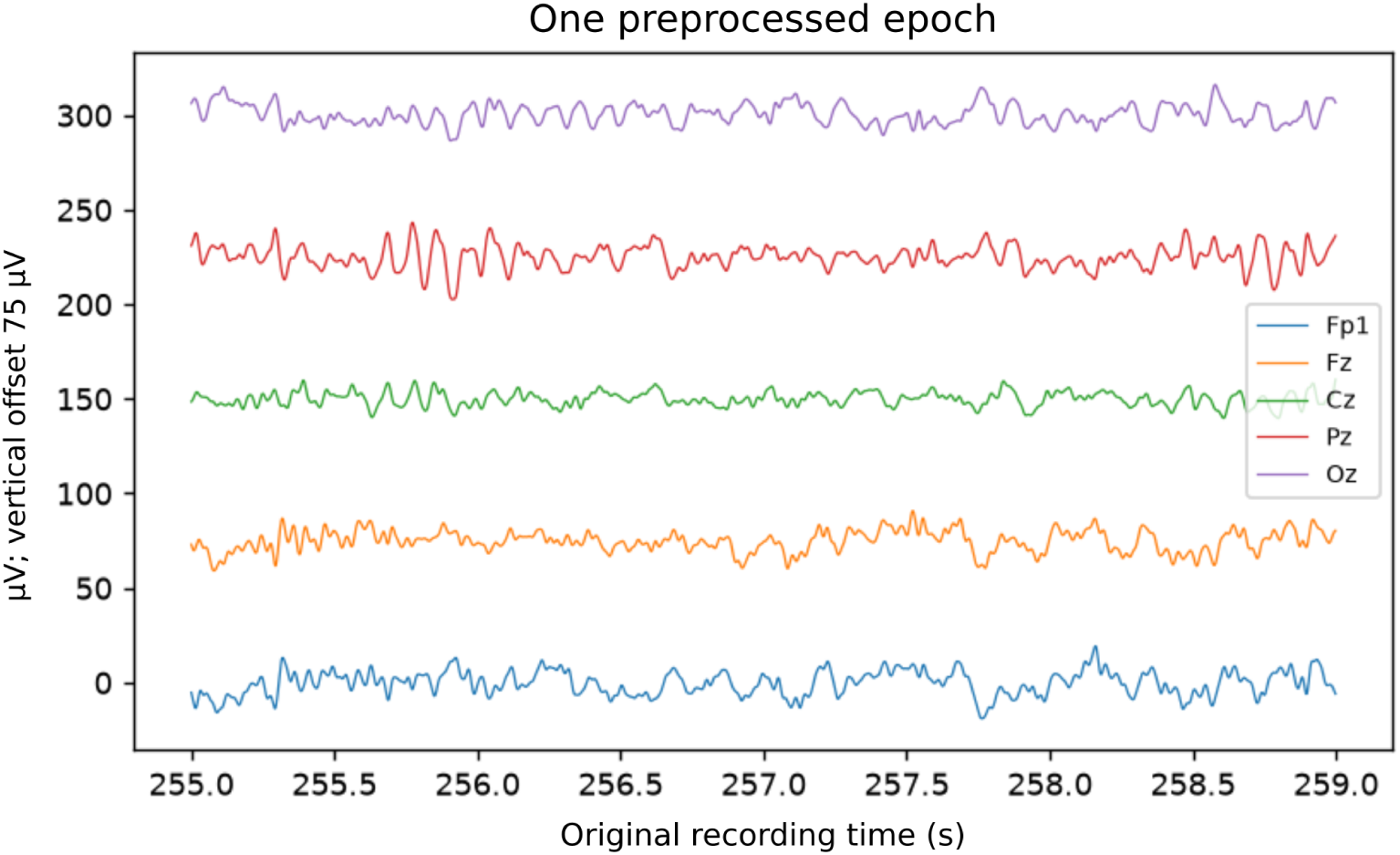
Individual example of preprocessed EEG at Fp1, Fz, Cz, Pz and Oz. The four-second epoch begins at second 255 of the original recording and has a sampling rate of 512 Hz. Time is in seconds and amplitude in *µ*V, with a visual vertical offset of 75 *µ*V.

Each trace is centred on its median and shifted vertically for readability; the vertical values include that offset. Fp1 is an additional illustrative channel outside the 14-position regional analysis. This individual example does not establish between-group differences.

## 3 Results

### 3.1 Participants and regional coverage

We considered 130 candidate recordings: 80 from exposed participants and 50 from controls. Power estimates were available for 122. The remaining eight had unavailable signal (one), channel labels whose correspondence to EEG positions was unconfirmed (six), or insufficient usable channels (one).

Each sensor core required all its positions, without imputation. A missing position affected only its corresponding core; available sample sizes therefore vary across regions (Table 2).

### 3.2 Absolute power and descriptive alpha measures

Table 4 reports median alpha power by sensor core. Figure 4 shows individual occipital alpha distributions. These summaries describe the observed data without covariate adjustment.

**Figure 3.**
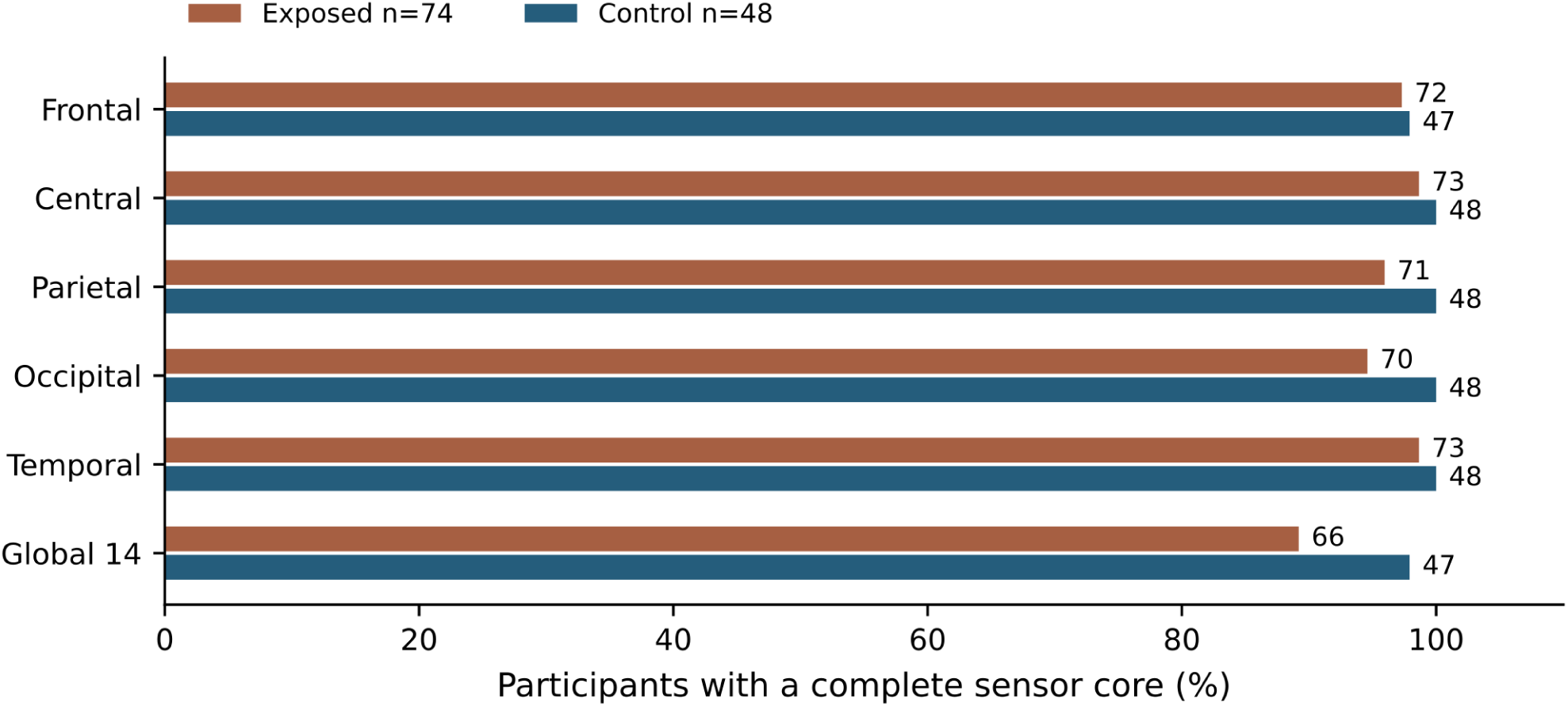
Participants with a complete sensor core. Numbers at the ends of the bars denote people; percentages use the power-available group as denominator: 74 exposed participants and 48 controls.

**Figure 4.**
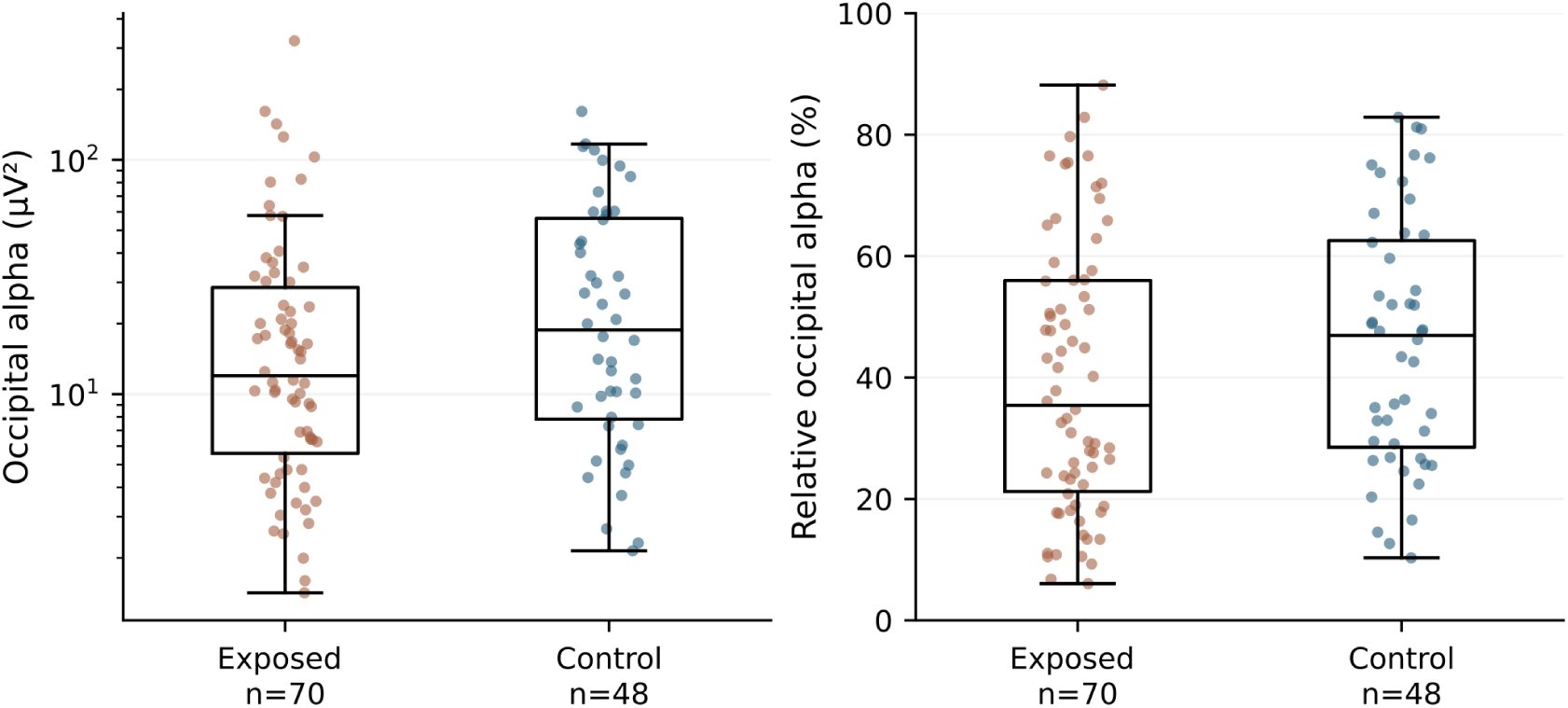
Absolute and relative occipital alpha power by participant. Each point represents one person. Boxes show the median and interquartile range; absolute power is displayed on a logarithmic scale. These are unadjusted descriptions.

**Table 4.**
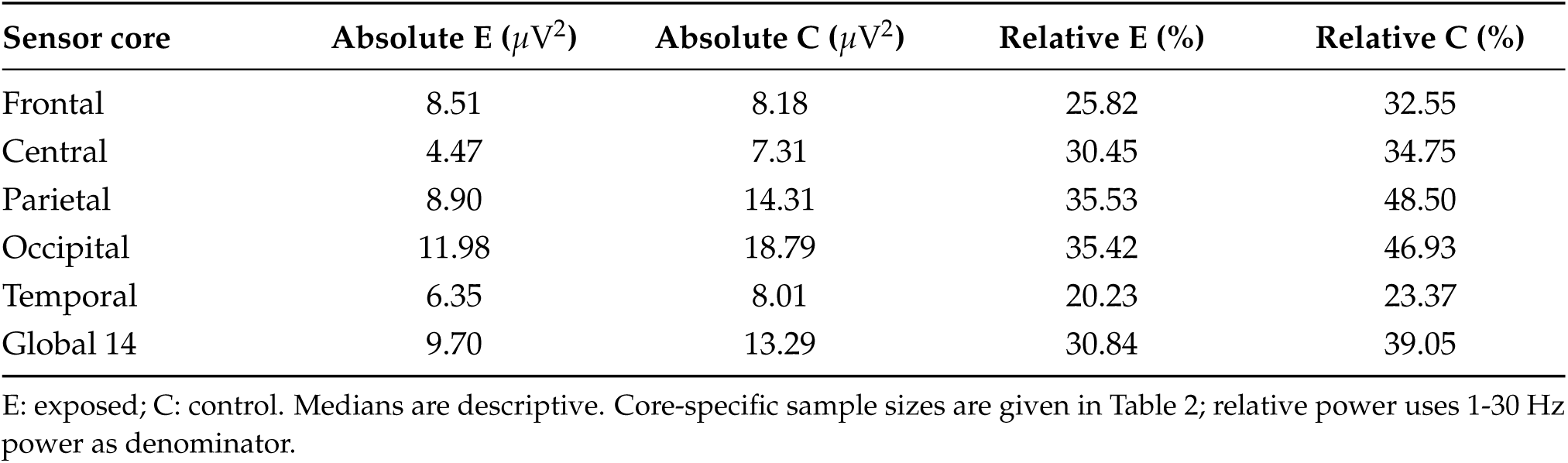
Median absolute and relative alpha power by sensor core.

Median absolute occipital alpha power was 11.98 *µ*V^2^ in exposed participants and 18.79 *µ*V^2^ in controls; relative-power medians were 35.42% and 46.93%. These values describe the observed distributions, while the models below quantify adjusted contrasts. Relative power expresses 1-30 Hz spectral composition and can change through shifts in other bands.

For occipital alpha, the adjusted exposed/control ratio was 0.87 (95% CI 0.45 to 1.68; 70 exposed participants and 46 controls). For the global 14-position core it was 0.94 (0.51 to 1.71; 66 and 45). Both intervals were compatible with the null value and with meaningful differences in either direction. Their overall Holm-adjusted values were 1.000.

### 3.3 Adjusted contrasts across four frequency bands

Figure 5 presents all 24 absolute-power contrasts. A ratio of 1 is the model’s null value; values below 1 correspond to lower power in exposed participants. Table 5 summarises the global-core absolute and relative effects across the four bands.

**Figure 5.**
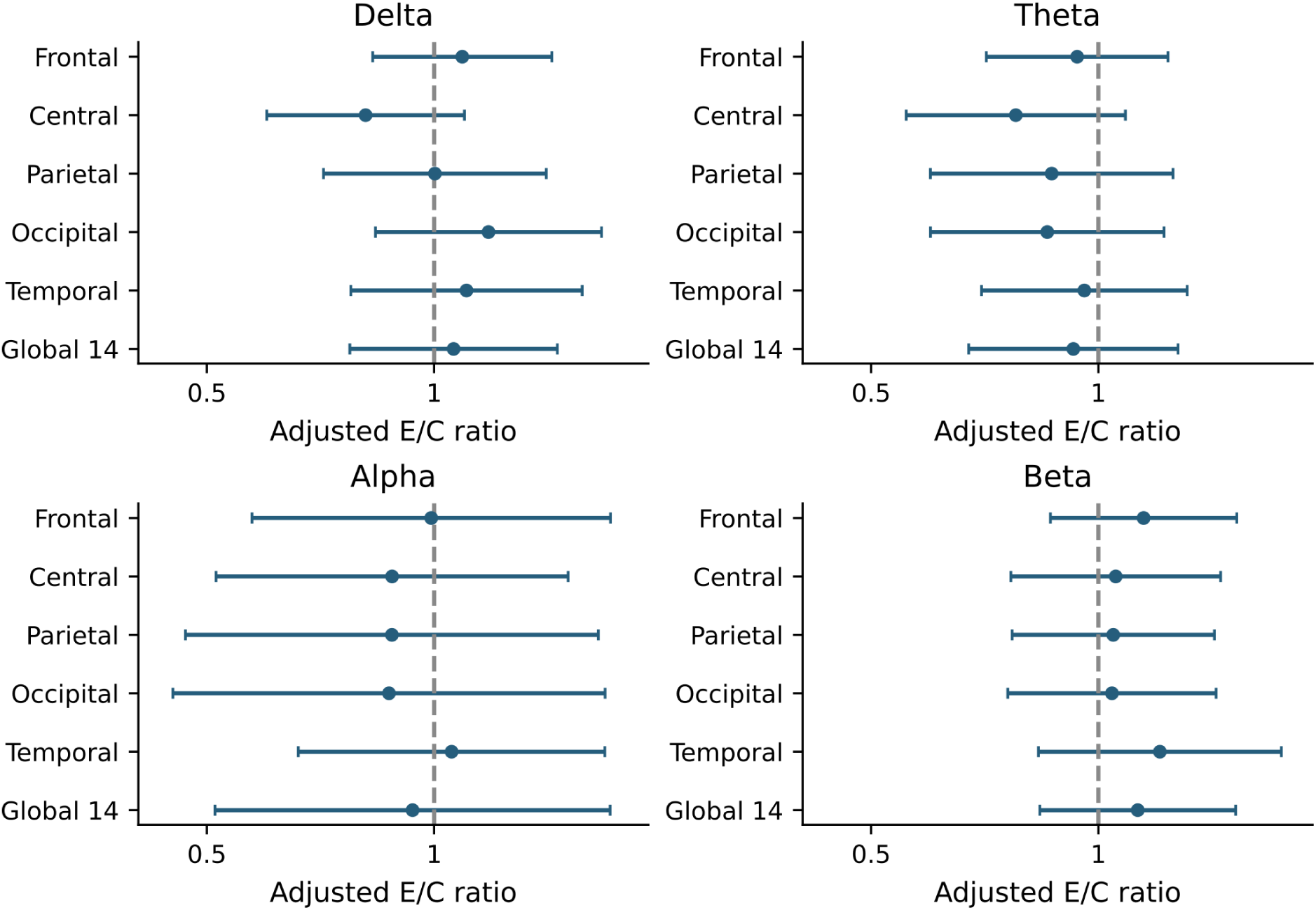
Adjusted exposed/control ratios with nominal 95% CIs. The line at 1 indicates the null ratio. Intervals are not simultaneous.

**Table 5.** Adjusted contrasts in the global 14-position core.

| Band | Absolute E/C ratio and 95% CI | Relative difference (pp) and 95% CI |
| --- | --- | --- |
| Delta | 1.06 (0.77 to 1.46) | 2.99 (−5.11 to 11.08) |
| Theta | 0.93 (0.67 to 1.27) | −1.75 (−4.57 to 1.08) |
| Alpha | 0.94 (0.51 to 1.71) | −1.93 (−13.04 to 9.19) |
| Beta | 1.13 (0.84 to 1.52) | 0.69 (−3.05 to 4.42) |

Both measures use 66 exposed participants and 45 controls. E/C: exposed/control ratio. pp: percentage points. The 95% confidence intervals (CIs) are nominal rather than simultaneous.

**Table 6.** Sensitivity of absolute alpha power to reference and assessment year.

| Sensor core | Reference or sample | n E/C | E/C ratio | 95% CI |
| --- | --- | --- | --- | --- |
| Occipital | EEG average | 70/46 | 0.87 | 0.45 to 1.68 |
| Occipital | Fz/Cz/Pz/Oz | 70/46 | 0.80 | 0.41 to 1.53 |
| Occipital | Common assessment year | 29/46 | 1.03 | 0.51 to 2.08 |
| Global 14 | EEG average | 66/45 | 0.94 | 0.51 to 1.71 |
| Global 14 | Fz/Cz/Pz/Oz | 66/45 | 0.93 | 0.51 to 1.71 |
| Global 14 | Common assessment year | 26/45 | 1.03 | 0.55 to 1.93 |

Average and fixed references are compared in the same participants. Common assessment year: average reference restricted to 2025; this restriction also changes sample composition.

The 78 EEG contrasts quantify adjusted magnitudes and their precision. None met the Holm-adjusted significance criterion, either within its family or across the planned set. Establishing equivalence would require a dedicated analysis.

### 3.4 Relative power and sensitivity

Figure 6 summarises adjusted relative-power differences in percentage points (pp). Colour indicates magnitude and direction rather than significance.

**Figure 6.**
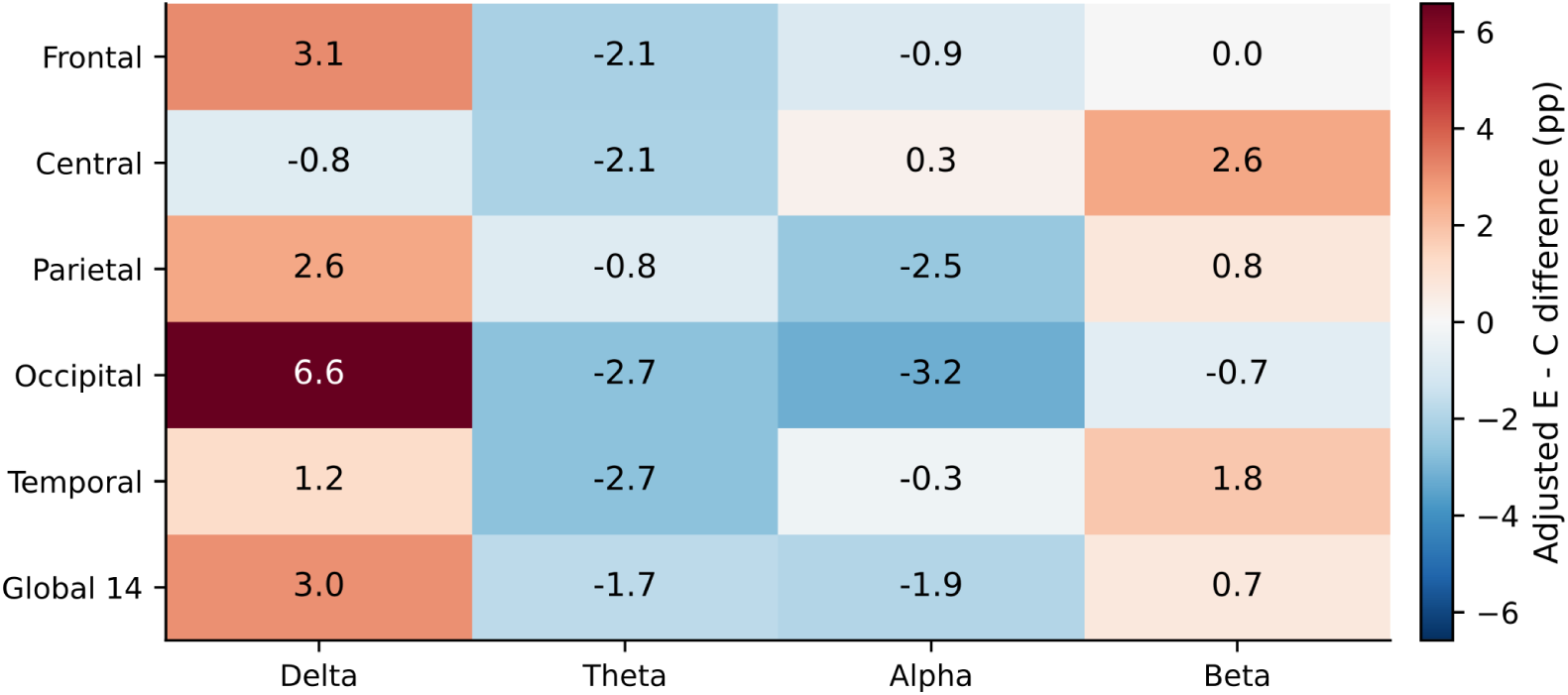
Adjusted relative-power differences in percentage points, exposed minus control. Colours describe magnitude and direction rather than statistical significance.

For temporal theta, the adjusted coefficient was *−*2.68 pp (95% CI *−*5.03 to *−*0.32; 73 exposed participants and 46 controls), with a nominal p value of 0.026. Within-family correction yielded p=0.626, and the overall correction yielded p=1.000. This result provides an exploratory observation for follow-up.

Under the fixed reference, ratios were 0.93 for global alpha and 0.80 for occipital alpha; restriction to 2025 yielded 1.03 for both. All intervals included 1. Reference comparisons used the same participants; year restriction changed sample composition and was not an independent confirmation.

### 3.5 Functional connectivity and matrices

Connectivity was estimated using wPLI in theta, alpha and beta over the 14-sensor core. Figure 7 presents descriptive group means and adjusted between-core differences. Diagonals are blank because self-connectivity was not estimated.

**Figure 7.**
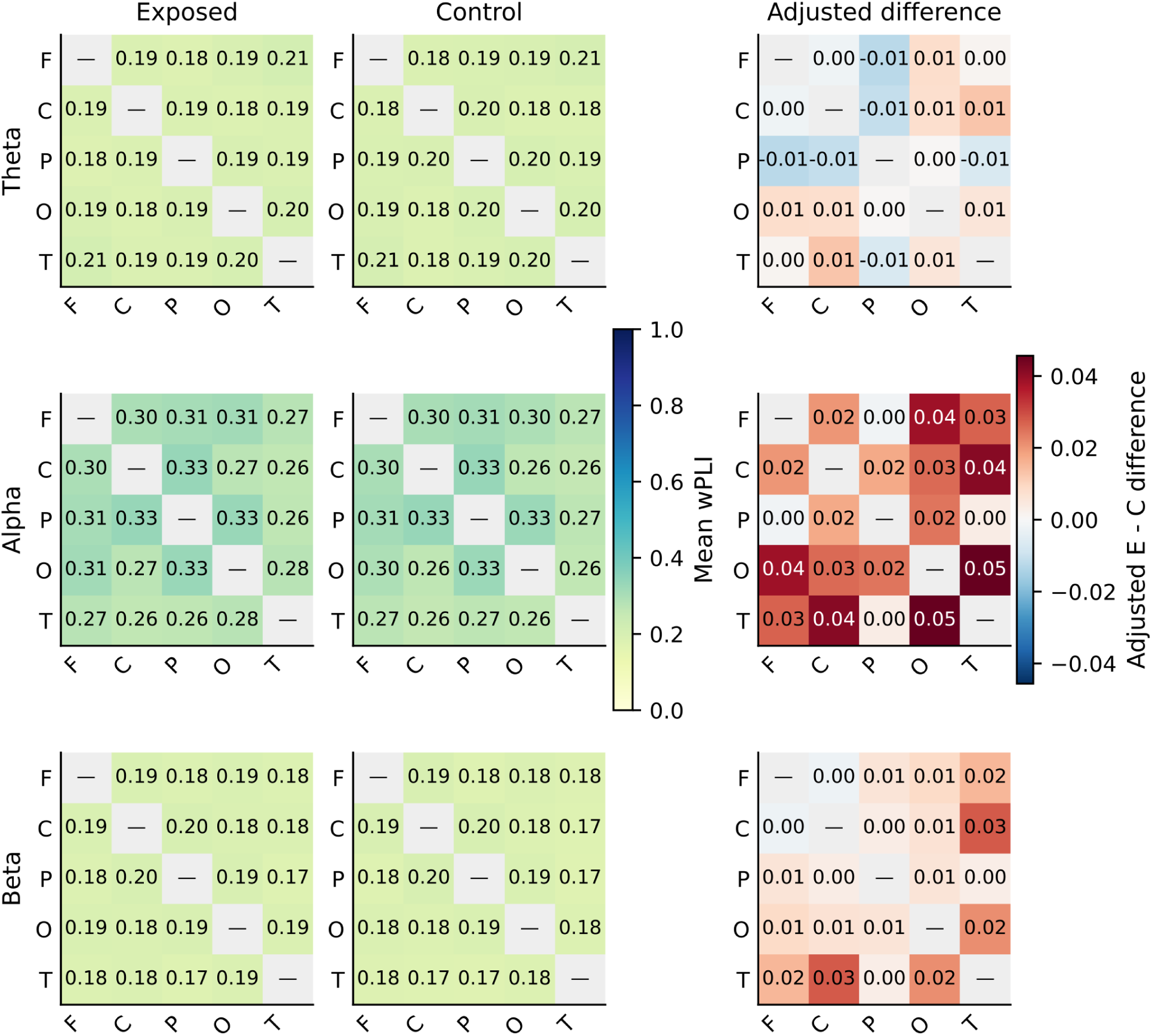
Regional connectivity matrices in theta, alpha and beta. F: frontal; C: central; P: parietal; O: occipital; T: temporal. The first two columns show descriptive participant means (66 exposed participants and 47 controls). The third shows adjusted differences among participants with complete covariates (66 and 45). Diagonals are unestimated; colour does not indicate significance.

Regional wPLI contrasts did not meet the corrected significance criterion. For alpha, the adjusted occipital-temporal difference was +0.046 (95% CI *−*0.002 to +0.094; 66 exposed participants and 45 controls). The interval included zero. Inference concerns the 30 regional-pair contrasts across the three bands, rather than each cell of the sensor maps.

#### 3.5.1 Alpha connectivity between sensors in exposed participants

Figure 8 shows the descriptive mean across the 14 positions in 66 exposed participants. It uses the same 0-1 scale as the control map to support direct visual comparison.

**Figure 8.**
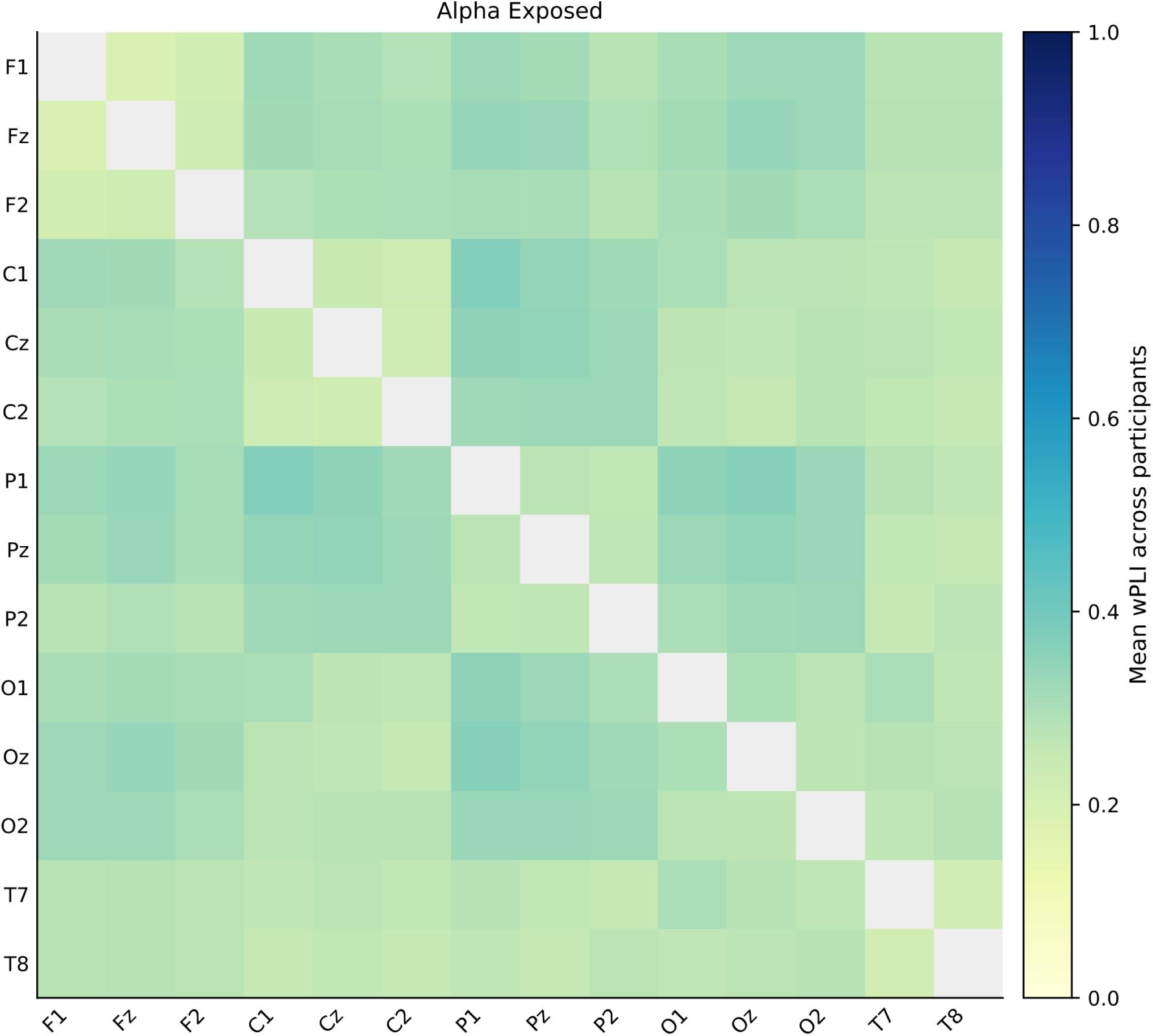
Mean alpha wPLI between sensors in exposed participants, n=66. The diagonal was not estimated. Colours and values are descriptive.

Shared activity also depends on the reference and on scalp signal mixing. This map describes sensor relationships rather than anatomical connections between brain regions.

#### 3.5.2 Alpha connectivity between sensors in controls

Figure 9 shows the descriptive mean over the same positions in 47 controls. The adjusted regional contrasts used 45 controls with complete covariates, explaining the difference between descriptive and inferential sample sizes.

**Figure 9.**
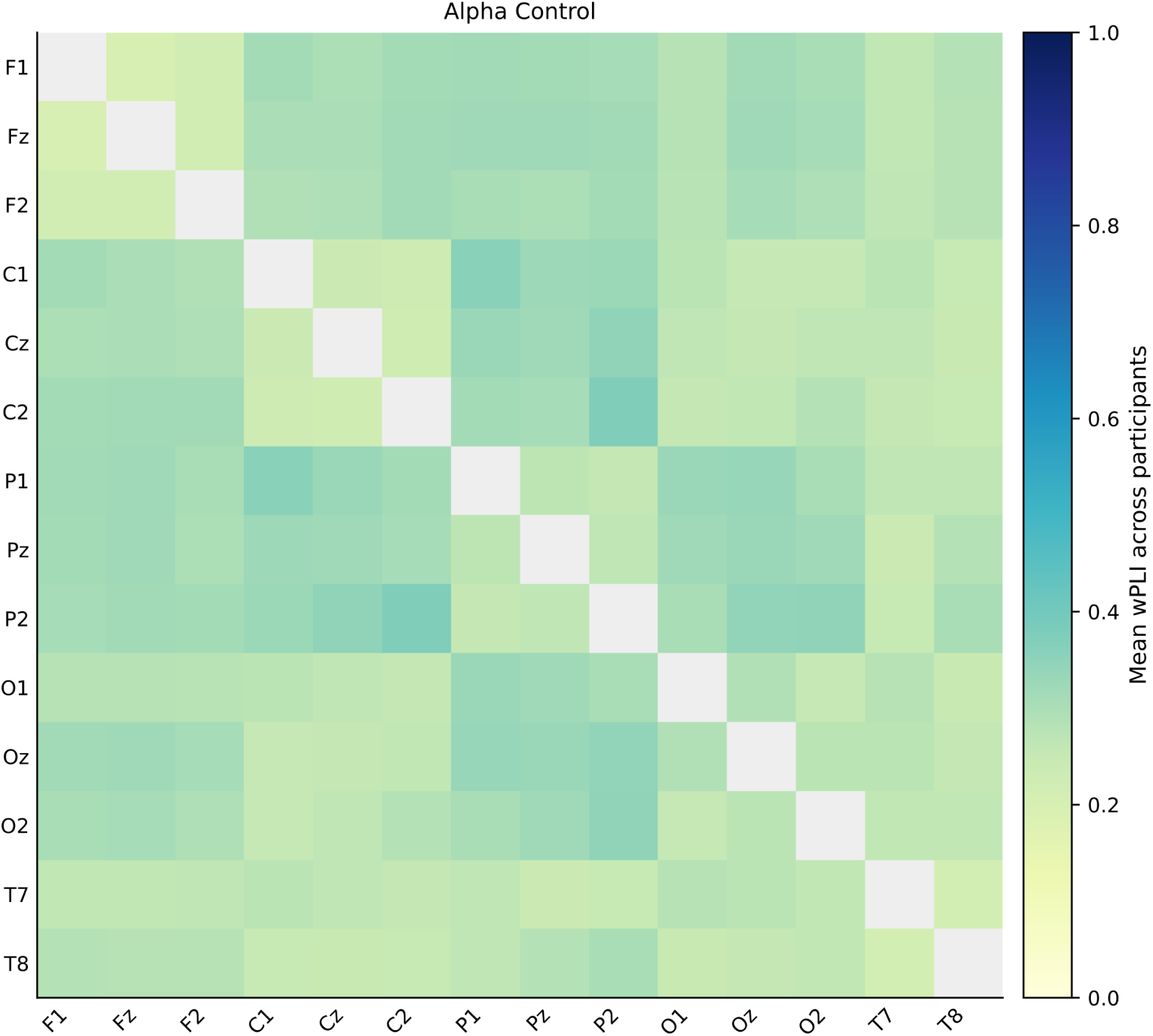
Mean alpha wPLI between sensors in controls, n=47. The 0-1 scale is shared with the exposed-group map. The diagonal was not estimated; colours and values are descriptive.

These maps describe the organisation of relationships between sensors. Group comparisons rely on the adjusted regional contrasts and their intervals in Figure 7 and Appendix C.

### 3.6 MoCA and its integration with EEG

MoCA adds cognitive performance information to EEG characterisation. It is integrated through a scoring review and three descriptive relationships, clarifying available information and supporting the planned cognitive comparison.

#### 3.6.1 Availability and scoring consistency

Of the 122 EEG participants, 120 have a reported MoCA total: 91 have no flags under the review rules and 29 require scoring confirmation. Table 7 and Figure 10 summarise the single exposed group and the control group.

**Figure 10.**
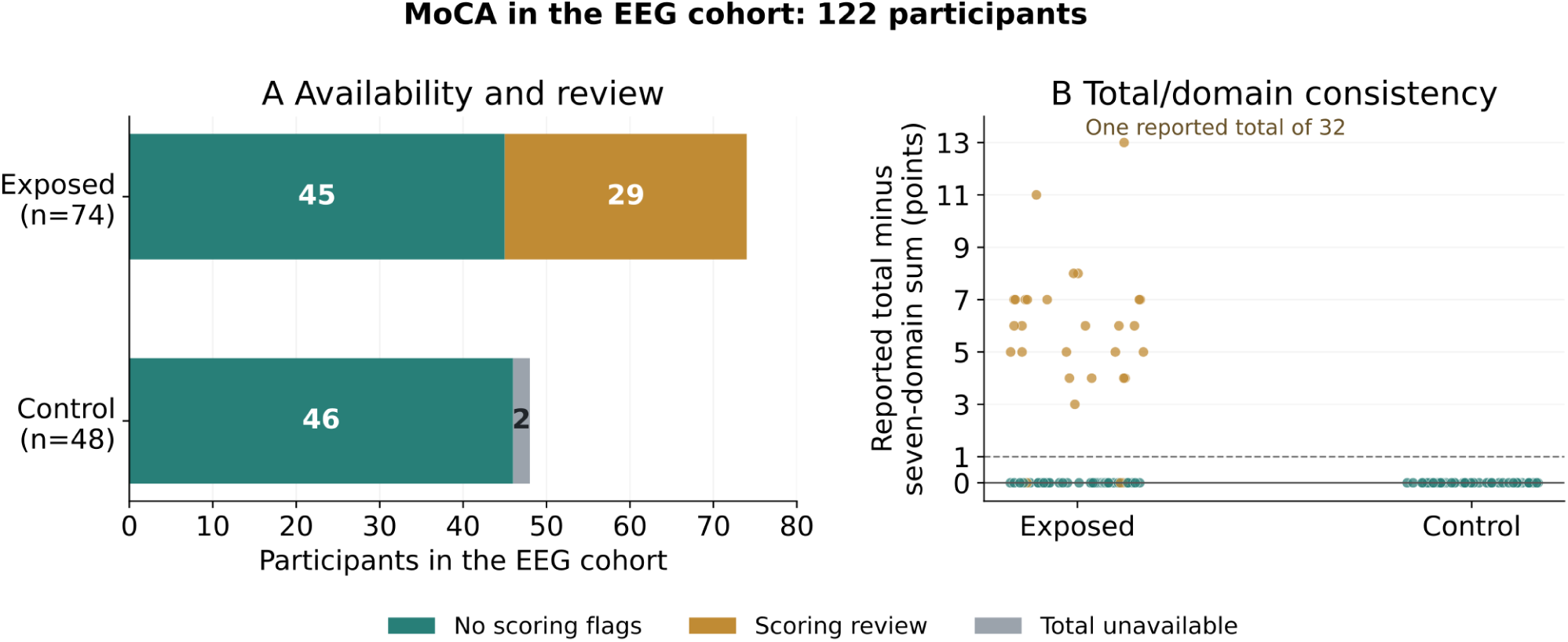
MoCA scoring review in the EEG cohort. A: availability and scoring-review status within the exposed and control groups. B: reported total minus the sum of seven domains in the 120 participants with a total. Lines at 0 and 1 are review references rather than applied corrections. Horizontal jitter separates overlapping observations.

**Figure 11.**
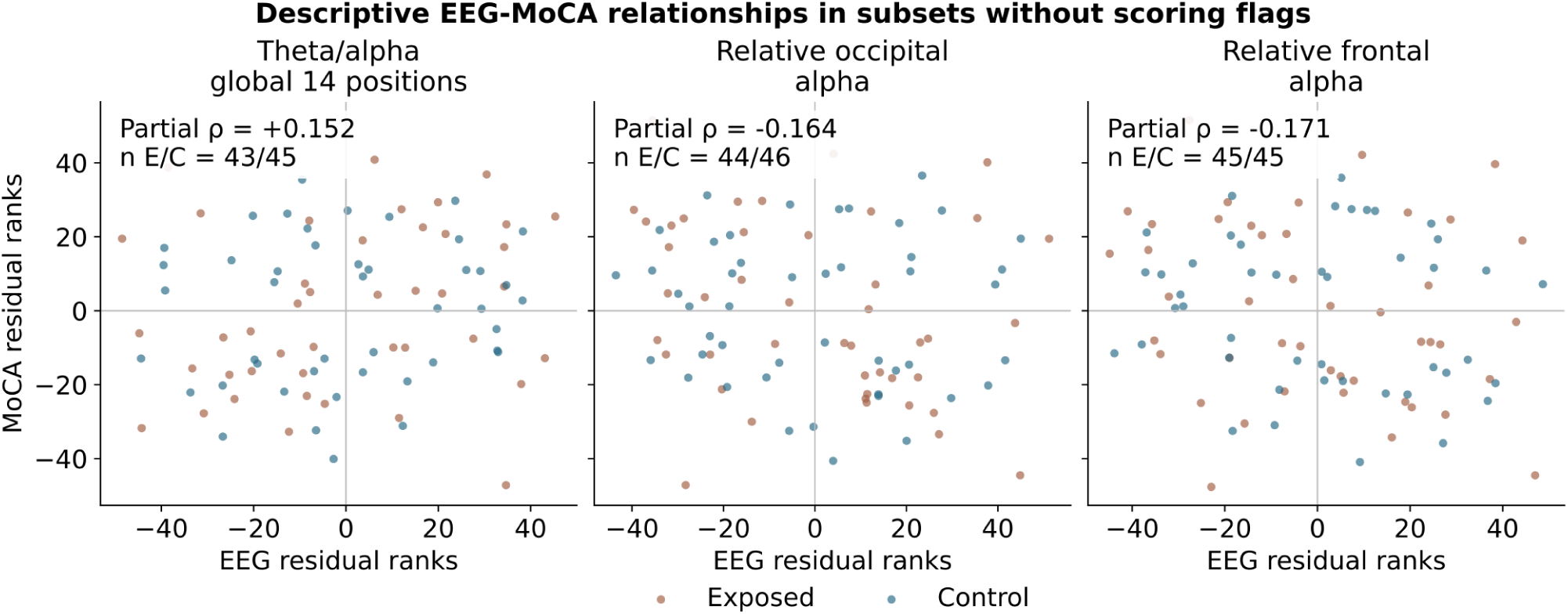
Descriptive EEG-MoCA rank relationships after covariate adjustment. Each point represents an eligible participant; colours identify the single exposed group and controls. The correlations describe the stated subsets and do not constitute a test of cognitive differences between groups.

**Table 7.** MoCA availability and scoring consistency in the EEG cohort.

| Group | EEG | Total available | No flags | Review |
| --- | --- | --- | --- | --- |
| Exposed | 74 | 74 | 45 | 29 |
| Control | 48 | 46 | 46 | 0 |
| Total | 122 | 120 | 91 | 29 |

Two controls have no available total or assessment year. No flags indicates consistency under the review rules; scoring validation requires confirmation of the test version and its instructions.

**Table 8.** Descriptive rank associations between EEG and total MoCA.

| EEG measure | n E/C | Total | Partial $\rho$ |
| --- | --- | --- | --- |
| Global 14-position theta/alpha | 43/45 | 88 | +0.152 |
| Relative occipital alpha | 44/46 | 90 | -0.164 |
| Relative frontal alpha | 45/45 | 90 | -0.171 |

E: exposed; C: control. Subsets have a reported total without scoring flags and complete EEG measures and covariates. Adjustment uses ranked age and education and categorical sex, assessment year and group. Coefficients are descriptive, without significance tests or CIs.

In 25 cases, the reported total exceeds the domain sum by 3-11 points; one further case has a total of 32 and a difference of 13. Three additional flags concern component scores.

#### 3.6.2 Descriptive relationships between EEG and MoCA

Global theta/alpha had a partial rank coefficient of 0.152; relative occipital and frontal alpha had coefficients of *−*0.164 and *−*0.171. Magnitudes below 0.18 characterise modest relationships and provide an initial description of variation shared between EEG and cognitive performance.

Eligible subsets are described in Table 8 and vary with the availability of complete EEG measures and covariates. All exposed participants were analysed as one pooled group. Interpretation of the coefficients is limited to these subsets. The four inferential MoCA models await scoring confirmation; these descriptive coefficients add no significance tests to the Holm set.

## 4 Discussion

We examined whether exposure to armed conflict was associated with differences in resting state EEG spectral power and functional connectivity in Colombian adults and explored the relationship between EEG activity and cognitive performance. Across 122 participants, including 74 exposed individuals and 48 controls, we evaluated absolute and relative power in delta, theta, alpha and beta frequencies and regional connectivity using wPLI, with adjustment for age, education, sex and assessment year. Descriptively, exposed participants showed lower occipital and global alpha power; the adjusted ratios were close to one and confidence intervals were wide. Temporal theta relative power was lower in exposed participants at nominal significance. None of the 78 EEG contrasts met the significance threshold after Holm correction. Exploratory EEG associations with MoCA performance were modest. Together, the estimates and their uncertainty provide a basis for refining hypotheses about resting EEG after conflict exposure and planning studies with greater precision.

The findings are relevant because the biological consequences of armed conflict remain considerably less studied than its psychiatric effects. Meta analytic evidence indicates that 22.1% of people living in conflict affected settings have a mental disorder at any point in time, including depression, anxiety, post traumatic stress disorder, bipolar disorder or schizophrenia [1]. In Colombia, conflict affected and displaced populations show substantial burdens of PTSD, depression, anxiety and somatic symptoms, and culturally adapted treatment can produce large improvements in these outcomes [6]. Our results extend this literature from psychiatric symptoms to direct measurement of brain function. The present comparisons do not establish a consistent group difference after correction, and the broad intervals remain compatible with effects of potential interest in either direction. This uncertainty creates a clear opportunity to examine whether individual trajectories, exposure characteristics, current psychopathology and social conditions account for variation that a single exposed/control indicator may not capture.

The alpha findings warrant particular consideration. Alpha oscillations are sensitive to arousal, attention and the organization of internally directed brain activity, and altered alpha activity has been reported across trauma related conditions. Systematic reviews of PTSD electrophysiology have repeatedly identified alpha rhythms among the EEG features associated with post traumatic symptoms [3]. In our study, exposed participants showed descriptively lower occipital and global alpha power. The adjusted exposed to control ratios were 0.87 and 0.94, respectively, with both confidence intervals spanning substantial effects in either direction. These estimates support further examination of alpha activity alongside demographic, clinical and exposure characteristics, while leaving its value as a marker of conflict exposure to be established in independent samples.

Theta activity showed a similarly nuanced pattern. Temporal relative theta was 2.68 percentage points lower in exposed participants and reached nominal significance. The contrast did not retain statistical significance after Holm correction. Previous PTSD studies have reported higher frontal and parietal theta power, including a resting EEG study of 17 patients with PTSD and 17 controls [4]. The difference in direction and anatomical distribution is important. Our sample was defined by exposure to armed conflict rather than by PTSD diagnosis, and lived exposure to conflict encompasses a much wider range of psychological responses, resilience trajectories and social circumstances. Neuroimaging meta analysis has similarly shown that trauma can influence brain function even among individuals without PTSD, while the transition from exposure to clinical disorder is associated with additional neural differences [11]. Future analyses that model trauma severity, PTSD symptoms, depression, anxiety and cumulative exposure continuously could clarify how these dimensions relate to EEG variation within and between the study groups.

The connectivity results also differ from some previous PTSD findings. Resting EEG studies of clinically diagnosed PTSD have reported altered alpha connectivity and reduced network strength, clustering and efficiency across theta, delta and beta frequencies [4, 5]. No regional wPLI contrast met the significance threshold after correction. The largest alpha estimate involved occipital temporal connectivity, with an exposed minus control difference of 0.046 and a confidence interval that included zero. The interval leaves the size of this contrast uncertain. Differences in population characteristics and in sensor level versus source based measures offer plausible questions for subsequent comparisons, although the present analysis cannot determine which accounts for the pattern. Connectivity findings in clinically selected PTSD samples therefore provide hypotheses to evaluate in broader conflict exposed populations.

The modest relationships between resting EEG and cognition provide another perspective on these findings. Global theta to alpha ratio showed a partial correlation of 0.152 with MoCA, while relative occipital and frontal alpha showed correlations of *−*0.164 and *−*0.171. These small effects suggest limited shared variation between the selected resting EEG measures and global cognitive performance in the current sample. Their interpretation remains preliminary because 29 MoCA scores require confirmation and inferential cognitive analyses remain pending. Resting EEG may also relate more strongly to specific cognitive processes than to a global screening score. Attention, episodic memory, executive control and emotional processing may therefore provide more sensitive phenotypes in subsequent analyses.

The Colombian context is central to interpreting these results. Armed conflict in Colombia has involved prolonged exposure to violence, displacement, social exclusion and material deprivation, with consequences extending across mental health, social functioning and brain health. Recent work has emphasized that more than seven million Colombians have been displaced and that violence interacts with poverty, disrupted communities and environmental degradation to generate a broader brain health challenge [7]. These exposures rarely occur in isolation. Age, education and assessment year already differed between groups in our sample, and other unmeasured dimensions such as childhood adversity, displacement history, socioeconomic conditions, ongoing insecurity, physical health and access to care may contribute to neurophysiological variability. Contemporary brain health frameworks increasingly emphasize resilience and the interaction between social, environmental and biological determinants across diverse populations [12]. This broader framework may be particularly appropriate for populations affected by chronic armed conflict.

Several features strengthen the study. EEG duration was standardized to 240 seconds per participant, with identical frequency definitions and common regional sensor cores across groups. Regional analyses required complete sensor sets and did not impute missing positions. Spectral and connectivity analyses covered complementary dimensions of neural activity, and group models incorporated age, education, sex and assessment year. Statistical inference used HC3 robust covariance and explicit correction across a large family of EEG comparisons. Sensitivity analyses tested alpha estimates under an alternative reference and after restricting participants to the common assessment year. The study also extends neurophysiological research into a Colombian population directly affected by armed conflict, a setting that remains poorly represented in biomarker research despite the scale and duration of exposure.

### 4.1 Limitations

Several limitations constrain interpretation. The sample provides limited power for small and spatially heterogeneous EEG effects, as reflected in the broad confidence intervals. The exposed and control groups differed in age, education and assessment year, and regression adjustment cannot fully remove residual confounding associated with these differences. Exposure to armed conflict was treated at the group level, without modelling the type, timing, duration or cumulative severity of violence, displacement and related adversities. Psychiatric symptoms were not incorporated into the present EEG models, limiting direct comparison with studies of PTSD. The analysis was performed at the sensor level and cannot localize cortical generators or establish anatomical connectivity. No ICA or interpolation was applied, and the planned standardized visual review of ocular, muscular and vigilance related artefacts remains to be completed. MoCA analyses also remain preliminary because 29 reported scores require verification. Finally, the cross sectional design cannot establish whether neurophysiological differences preceded exposure, emerged following conflict related experiences or reflect subsequent social and health trajectories.

## 5 Conclusion

This study characterized resting EEG in 122 Colombian adults using a common analysed duration, regional sensor cores and adjusted comparisons. None of the 78 EEG contrasts met the significance threshold after Holm correction; the confidence intervals leave several effect magnitudes uncertain. Descriptive alpha patterns, the temporal theta estimate and exploratory EEG-MoCA relationships provide specific questions for follow-up, with cognitive inference remaining conditional on scoring confirmation. Larger longitudinal studies that integrate detailed histories of violence and displacement with psychiatric, cognitive and social measures could clarify when conflict exposure is associated with neurophysiological variation. The present findings provide effect estimates and a reproducible analytical framework to guide that work.

## Author contributions

LN-P, SB, PR and HS-G conceived the proposal. LN-P and HS-G wrote the drafts. SB and LN- P prepared the analyses, tables and figures under the supervision of HS-G and PR. SB and LN-P implemented the EEG analyses. HS-G coordinated specific manuscript sections. All authors reviewed, edited, provided critical comments, and approved the manuscript before submission.

## Data Availability

The data are not publicly available because they were collected from people in a highly vulnerable situation, and sharing them could compromise their privacy and safety.

## Footnotes

1 MoCA Cognition. Version-specific scoring instructions: https://mocacognition.com/faq/.

## A Absolute power comparisons

Table 9 includes all 24 adjusted contrasts. E: exposed; C: control; n: participants with complete cases. The effect is the E/C ratio of conditional geometric means, with a null value of 1. CIs are nominal. The Holm p column refers to the planned set of 82 contrasts described in Section 2.

**Table 9.**
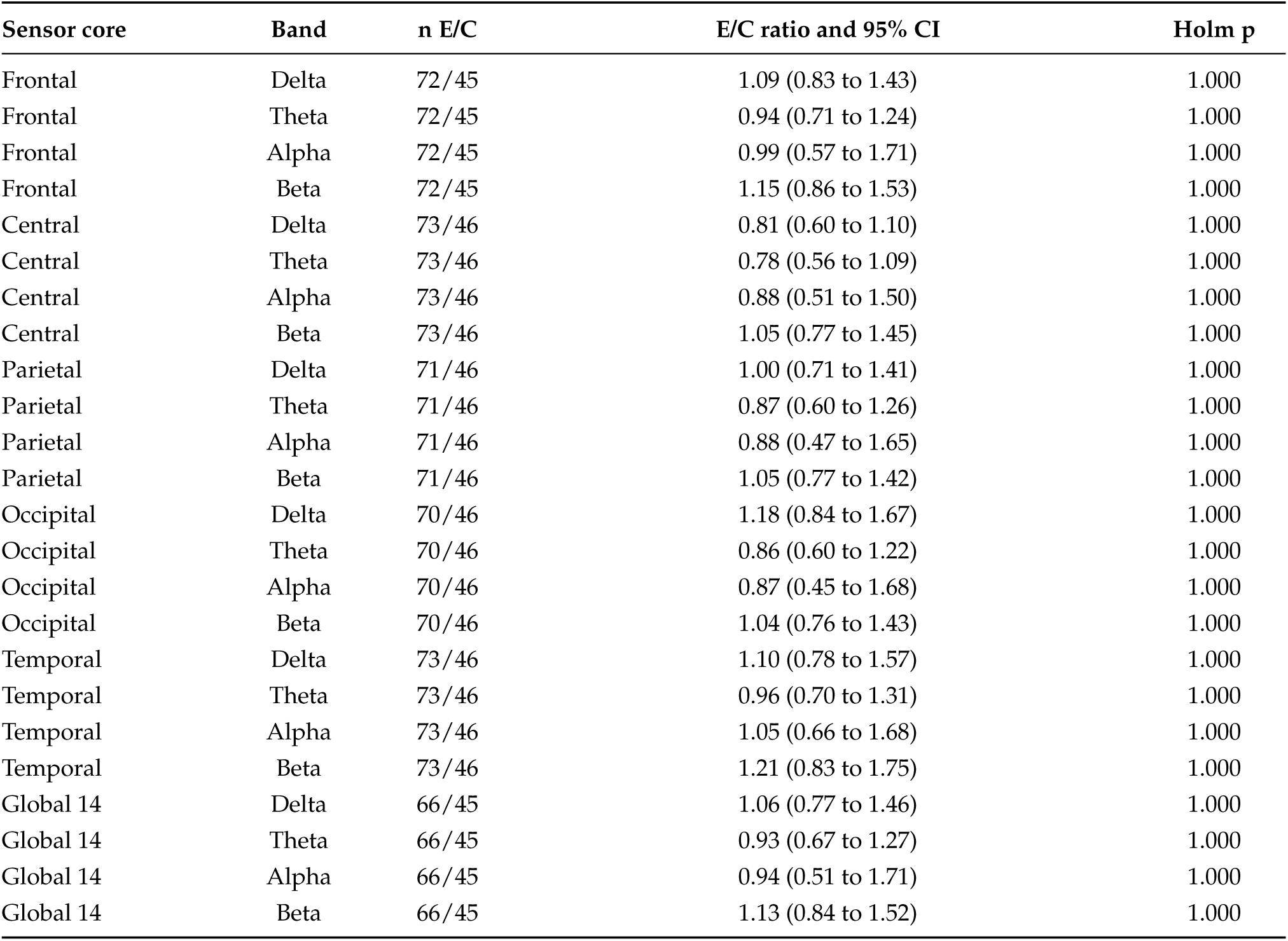
Adjusted absolute-power comparisons by band and sensor core.

## **B** Relative power comparisons

Table 10 includes all 24 adjusted contrasts. Effects are exposed-minus-control differences in percentage points, with a null value of 0. E: exposed; C: control. CIs are nominal. The Holm p column refers to the planned set of 82 contrasts.

**Table 10.** Adjusted relative-power comparisons by band and sensor core.

| Sensor core | Band | n E/C | Difference (pp) and 95% CI | Holm p |
| --- | --- | --- | --- | --- |
| Frontal | Delta | 72/45 | 3.10 (−4.31 to 10.51) | 1.000 |
| Frontal | Theta | 72/45 | −2.12 (−5.57 to 1.33) | 1.000 |
| Frontal | Alpha | 72/45 | −0.94 (−11.36 to 9.48) | 1.000 |
| Frontal | Beta | 72/45 | −0.04 (−3.42 to 3.33) | 1.000 |
| Central | Delta | 73/46 | −0.77 (−7.72 to 6.17) | 1.000 |
| Central | Theta | 73/46 | −2.07 (−4.72 to 0.59) | 1.000 |
| Central | Alpha | 73/46 | 0.27 (−8.87 to 9.42) | 1.000 |
| Central | Beta | 73/46 | 2.57 (−0.52 to 5.66) | 1.000 |
| Parietal | Delta | 71/46 | 2.56 (−5.69 to 10.81) | 1.000 |
| Parietal | Theta | 71/46 | −0.83 (−4.18 to 2.52) | 1.000 |
| Parietal | Alpha | 71/46 | −2.49 (−14.19 to 9.21) | 1.000 |
| Parietal | Beta | 71/46 | 0.77 (−2.29 to 3.82) | 1.000 |
| Occipital | Delta | 70/46 | 6.58 (−2.12 to 15.28) | 1.000 |
| Occipital | Theta | 70/46 | −2.69 (−5.72 to 0.34) | 1.000 |
| Occipital | Alpha | 70/46 | −3.20 (−15.30 to 8.90) | 1.000 |
| Occipital | Beta | 70/46 | −0.70 (−5.08 to 3.69) | 1.000 |
| Temporal | Delta | 73/46 | 1.19 (−6.21 to 8.58) | 1.000 |
| Temporal | Theta | 73/46 | −2.68 (−5.03 to −0.32) | 1.000 |
| Temporal | Alpha | 73/46 | −0.28 (−8.01 to 7.44) | 1.000 |
| Temporal | Beta | 73/46 | 1.77 (−4.65 to 8.19) | 1.000 |
| Global 14 | Delta | 66/45 | 2.99 (−5.11 to 11.08) | 1.000 |
| Global 14 | Theta | 66/45 | −1.75 (−4.57 to 1.08) | 1.000 |
| Global 14 | Alpha | 66/45 | −1.93 (−13.04 to 9.19) | 1.000 |
| Global 14 | Beta | 66/45 | 0.69 (−3.05 to 4.42) | 1.000 |

## **C** Regional connectivity comparisons

Table 11 includes all 30 adjusted contrasts. Effects are exposed-minus-control differences in wPLI, with a null value of 0. E: exposed; C: control. CIs are nominal. The Holm p column refers to the planned set of 82 contrasts. Five decimal places are retained in the central-temporal beta interval to preserve the sign of its lower bound near zero.

**Table 11.** Adjusted regional-connectivity comparisons by band.

| Sensor-core pair | Band | n E/C | wPLI difference and 95% CI | Holm p |
| --- | --- | --- | --- | --- |
| Frontal and central | Theta | 66/45 | −0.002 (−0.026 to 0.022) | 1.000 |
| Frontal and central | Alpha | 66/45 | 0.020 (−0.050 to 0.090) | 1.000 |
| Frontal and central | Beta | 66/45 | −0.002 (−0.020 to 0.015) | 1.000 |
| Frontal and parietal | Theta | 66/45 | −0.012 (−0.037 to 0.012) | 1.000 |
| Frontal and parietal | Alpha | 66/45 | −0.002 (−0.056 to 0.053) | 1.000 |
| Frontal and parietal | Beta | 66/45 | 0.006 (−0.005 to 0.017) | 1.000 |
| Frontal and occipital | Theta | 66/45 | 0.008 (−0.022 to 0.038) | 1.000 |
| Frontal and occipital | Alpha | 66/45 | 0.039 (−0.031 to 0.109) | 1.000 |
| Frontal and occipital | Beta | 66/45 | 0.009 (−0.008 to 0.025) | 1.000 |
| Frontal and temporal | Theta | 66/45 | 0.001 (−0.024 to 0.027) | 1.000 |
| Frontal and temporal | Alpha | 66/45 | 0.027 (−0.022 to 0.076) | 1.000 |
| Frontal and temporal | Beta | 66/45 | 0.015 (−0.017 to 0.047) | 1.000 |
| Central and parietal | Theta | 66/45 | −0.011 (−0.036 to 0.014) | 1.000 |
| Central and parietal | Alpha | 66/45 | 0.017 (−0.060 to 0.095) | 1.000 |
| Central and parietal | Beta | 66/45 | 0.003 (−0.016 to 0.023) | 1.000 |
| Central and occipital | Theta | 66/45 | 0.008 (−0.011 to 0.026) | 1.000 |
| Central and occipital | Alpha | 66/45 | 0.026 (−0.011 to 0.063) | 1.000 |
| Central and occipital | Beta | 66/45 | 0.007 (−0.010 to 0.023) | 1.000 |
| Central and temporal | Theta | 66/45 | 0.013 (−0.008 to 0.033) | 1.000 |
| Central and temporal | Alpha | 66/45 | 0.042 (−0.010 to 0.093) | 1.000 |
| Central and temporal | Beta | 66/45 | 0.028 (−0.00003 to 0.05633) | 1.000 |
| Parietal and occipital | Theta | 66/45 | 0.001 (−0.033 to 0.034) | 1.000 |
| Parietal and occipital | Alpha | 66/45 | 0.025 (−0.052 to 0.101) | 1.000 |
| Parietal and occipital | Beta | 66/45 | 0.006 (−0.012 to 0.024) | 1.000 |
| Parietal and temporal | Theta | 66/45 | −0.007 (−0.026 to 0.013) | 1.000 |
| Parietal and temporal | Alpha | 66/45 | 0.003 (−0.041 to 0.047) | 1.000 |
| Parietal and temporal | Beta | 66/45 | 0.003 (−0.009 to 0.015) | 1.000 |
| Occipital and temporal | Theta | 66/45 | 0.005 (−0.022 to 0.033) | 1.000 |
| Occipital and temporal | Alpha | 66/45 | 0.046 (−0.002 to 0.094) | 1.000 |
| Occipital and temporal | Beta | 66/45 | 0.021 (−0.014 to 0.056) | 1.000 |

